## Supplementary File for "MVMRmode: Introducing an R package for plurality valid estimators for multivariable Mendelian randomisation"

| Exposure 1 | Exposure 2 |
| --- | --- |
| 197 | 186 |

Supplementary Table 1: mean F statistic of the variant-exposure association for each setting over the 1000 simulations.

|  | | | No bias | 10% balanced pleiotropic | 20% balanced pleiotropic | 40% balanced pleiotropic | 10% directional pleiotropic | 20% directional pleiotropic | 40% directional pleiotropic |
| --- | --- | --- | --- | --- | --- | --- | --- | --- | --- |
| SD | IVW | Exposure 1 | 0.025 | 0.084 | 0.12 | 0.167 | 0.084 | 0.123 | 0.171 |
|  |  | Exposure 2 | 0.025 | 0.087 | 0.118 | 0.165 | 0.085 | 0.122 | 0.17 |
|  | MR Egger | Exposure 1 | 0.089 | 0.139 | 0.186 | 0.249 | 0.144 | 0.186 | 0.244 |
|  |  | Exposure 2 | 0.049 | 0.125 | 0.167 | 0.216 | 0.125 | 0.169 | 0.229 |
|  | Median | Exposure 1 | 0.031 | 0.035 | 0.041 | 0.05 | 0.035 | 0.039 | 0.051 |
|  |  | Exposure 2 | 0.032 | 0.037 | 0.041 | 0.05 | 0.036 | 0.039 | 0.052 |
|  | multivariable-CM | Exposure 1 | 0.028 | 0.03 | 0.033 | 0.044 | 0.031 | 0.042 | 0.108 |
|  |  | Exposure 2 | 0.028 | 0.03 | 0.034 | 0.044 | 0.03 | 0.039 | 0.103 |
|  | multivariable-MBE | Exposure 1 | 1.026 | 2.126 | 1.813 | 5.014 | 3.448 | 3.507 | 9.546 |
|  |  | Exposure 2 | 3.708 | 2.04 | 2.882 | 11.9 | 3.704 | 3.413 | 4.021 |
| Coverage | IVW | Exposure 1 | 0.948 | 0.946 | 0.945 | 0.951 | 0.945 | 0.932 | 0.904 |
|  |  | Exposure 2 | 0.941 | 0.938 | 0.942 | 0.944 | 0.934 | 0.908 | 0.878 |
|  | MR Egger | Exposure 1 | 0.639 | 0.873 | 0.892 | 0.918 | 0.873 | 0.904 | 0.938 |
|  |  | Exposure 2 | 0.828 | 0.924 | 0.943 | 0.958 | 0.925 | 0.935 | 0.945 |
|  | Median | Exposure 1 | 0.974 | 0.964 | 0.94 | 0.926 | 0.962 | 0.956 | 0.908 |
|  |  | Exposure 2 | 0.963 | 0.949 | 0.926 | 0.921 | 0.939 | 0.924 | 0.873 |
|  | multivariable-CM | Exposure 1 | 0.909 | 0.916 | 0.923 | 0.907 | 0.919 | 0.884 | 0.692 |
|  |  | Exposure 2 | 0.894 | 0.904 | 0.91 | 0.903 | 0.921 | 0.925 | 0.739 |
|  | multivariable-MBE | Exposure 1 | 0.995 | 0.994 | 0.996 | 0.995 | 0.991 | 0.993 | 0.977 |
|  |  | Exposure 2 | 0.991 | 0.991 | 0.995 | 0.992 | 0.992 | 0.996 | 0.986 |

Supplementary Table 2: Results for additional outcomes when both exposures cause the outcome, and exposure 2 is pleiotropic.

|  | | | No bias | 20% balanced pleiotropic | 40% balanced pleiotropic | 60% balanced pleiotropic | 10% directional pleiotropic | 20% directional pleiotropic | 40% directional pleiotropic |
| --- | --- | --- | --- | --- | --- | --- | --- | --- | --- |
| SD | IVW | Exposure 1 | 0.008 | 0.079 | 0.117 | 0.167 | 0.09 | 0.118 | 0.173 |
|  |  | Exposure 2 | 0.008 | 0.079 | 0.116 | 0.166 | 0.091 | 0.118 | 0.167 |
|  | MR Egger | Exposure 1 | 0.011 | 0.111 | 0.162 | 0.23 | 0.123 | 0.163 | 0.232 |
|  |  | Exposure 2 | 0.011 | 0.109 | 0.158 | 0.217 | 0.12 | 0.158 | 0.229 |
|  | Median | Exposure 1 | 0.01 | 0.012 | 0.014 | 0.02 | 0.012 | 0.014 | 0.021 |
|  |  | Exposure 2 | 0.01 | 0.012 | 0.014 | 0.021 | 0.012 | 0.014 | 0.021 |
|  | multivariable-CM | Exposure 1 | 0.008 | 0.012 | 0.021 | 0.043 | 0.024 | 0.064 | 0.134 |
|  |  | Exposure 2 | 0.008 | 0.013 | 0.021 | 0.045 | 0.024 | 0.064 | 0.143 |
|  | multivariable-MBE | Exposure 1 | 0.268 | 1.379 | 4.344 | 7.928 | 1.974 | 4.892 | 3.578 |
|  |  | Exposure 2 | 0.411 | 2.459 | 2.76 | 16.764 | 1.667 | 2.962 | 16.847 |
| Coverage | IVW | Exposure 1 | 0.957 | 0.956 | 0.947 | 0.957 | 0.935 | 0.931 | 0.902 |
|  |  | Exposure 2 | 0.946 | 0.962 | 0.951 | 0.945 | 0.922 | 0.92 | 0.883 |
|  | MR Egger | Exposure 1 | 0.948 | 0.953 | 0.945 | 0.954 | 0.94 | 0.963 | 0.95 |
|  |  | Exposure 2 | 0.951 | 0.949 | 0.944 | 0.947 | 0.944 | 0.958 | 0.948 |
|  | Median | Exposure 1 | 0.971 | 0.98 | 0.957 | 0.919 | 0.968 | 0.965 | 0.891 |
|  |  | Exposure 2 | 0.972 | 0.963 | 0.954 | 0.911 | 0.972 | 0.959 | 0.907 |
|  | multivariable-CM | Exposure 1 | 0.954 | 0.953 | 0.921 | 0.861 | 0.897 | 0.633 | 0.235 |
|  |  | Exposure 2 | 0.947 | 0.951 | 0.917 | 0.854 | 0.881 | 0.65 | 0.247 |
|  | multivariable-MBE | Exposure 1 | 0.994 | 0.992 | 0.992 | 0.99 | 0.992 | 0.987 | 0.989 |
|  |  | Exposure 2 | 0.997 | 0.995 | 0.99 | 0.989 | 0.991 | 0.99 | 0.98 |

Supplementary Table 3: Results for additional outcomes when neither exposure cause the outcome, and exposure 2 is pleiotropic.
